# Predicting dengue importation into Europe, using machine learning and model-agnostic methods

**DOI:** 10.1101/19013383

**Authors:** Donald Salami, Carla Alexandra Sousa, Maria do Rosário Oliveira Martins, César Capinha

## Abstract

The geographical spread of dengue is a global public health concern. This is largely mediated by the importation of dengue from endemic to non-endemic areas via the increasing connectivity of the global air transport network. The dynamic nature and intrinsic heterogeneity of the air transport network make it challenging to predict dengue importation.

Here, we explore the capabilities of state-of-the-art machine learning algorithms to predict dengue importation. We trained four machine learning classifiers algorithms, using a 6-year historical dengue importation data for 21 countries in Europe and connectivity indices mediating importation and air transport network centrality measures. Predictive performance for the classifiers was evaluated using the area under the receiving operating characteristic curve, sensitivity, and specificity measures. Finally, we applied practical model-agnostic methods, to provide an in-depth explanation of our optimal model’s predictions on a global and local scale.

Our best performing model achieved high predictive accuracy, with an area under the receiver operating characteristic score of 0.94 and a maximized sensitivity score of 0.88. The predictor variables identified as most important were the source country’s dengue incidence rate, population size, and volume of air passengers. Network centrality measures, describing the positioning of European countries within the air travel network, were also influential to the predictions.

We demonstrated the high predictive performance of a machine learning model in predicting dengue importation and the utility of the model-agnostic methods to offer a comprehensive understanding of the reasons behind the predictions. Similar approaches can be utilized in the development of an operational early warning surveillance system for dengue importation.

## Introduction

The geographical spread of dengue fever is a global public health concern. This spread, particularly to non-endemic areas, has been largely facilitated by an increase in global trade and human mobility^1-3^. The expansion and connectivity of the global air transport networks in recent years, has played a key role in this spread^3^. In Europe, where dengue is not endemic, the number of travel-related cases of dengue, demonstrates how the air transport network has facilitated the spread of the disease. In the past decade, the European region has reported a significant number of imported dengue cases from epidemic/endemic tropical and subtropical countries^4^. Sporadic autochthonous transmissions have also been triggered by imported cases in areas with suitable environmental conditions and an established presence of the mosquito vector^5^. Recent examples include the autochthonous cases reported in France and Spain, which were linked to having originated from an imported case^6^.

The mitigation of the continuous spread of dengue in Europe lies in part in the ability to effectively predict importation risk. However, a notable challenge in achieving this, is the complexity of global air transport networks, due to the dynamic nature and heterogeneity underlying the connections^1,7,8^. In recent times a range of modelling approaches, from the field of social network analysis, have been applied to understand the connection topology of the air transport network and their role in disease importation^9^. Unlike conventional statistical modelling approaches, these methods account for the co-dynamics of the network structure and how they interact with other risk factors to mediate the importation of dengue^10-12^. Our previous work^13^ integrates this modelling approach and offers a foundational understanding of the importation patterns of dengue in Europe.

Conversely, an increasing number of studies are employing the use of machine learning algorithms to develop robust predictive models for dengue^14,15^. Machine learning algorithms are an applied extension of artificial intelligence. These algorithms build a mathematical model base, to automatically learn data patterns, adjust and perform inference, without explicit instructions^16^. Several studies have demonstrated the powerful predictive capabilities of machine learning models and their superiority over conventional statistical methods^17-19^. To this effect, some studies have applied them in the development of predictive models for dengue incidence^14,20-22^. A recent study by Chen et al ^15^, utilized machine learning algorithms to develop a real-time model to forecast dengue in Singapore. Despite the high predictive performance of machine learning algorithms, they are not widely popular in epidemiological studies. This is likely to be in part because they are considered, to be “black-box” models with low interpretability, due to their complex inner workings. We argue different, that though machine learning models could fit complex relationships, several recent advancements have been made to aid the interpretation of these models^23^. To the best of our knowledge, machine learning algorithms have not been applied in modelling the risk of dengue importation for Europe.

Here, we aim to apply machine learning algorithms to develop a predictive model for dengue importation risk in Europe. To do so, we train a diverse set of machine learning algorithms, with historical data of dengue importation into Europe, connectivity indices of factors potentially mediating importation risk and centrality measures characterizing the air transport network. We then evaluate the predictive performance of the different models on a hold-out dataset to determine an optimal model. Finally, we employ the use of practical model-agnostic methods to interpret the optimal model’s predictions.

## Methods

### Dengue data

We obtained monthly data for imported cases of dengue in Europe, for 2010 – 2015, from the European Centre for Disease Prevention and Control (ECDC)^24^. Here, we utilized confirmed dengue cases (as defined the European Union generic case definition for viral haemorrhagic fevers) with known travel history^25^. A total of 21 European Union/European Economic Area (EU/EEA) countries reported data on imported dengue, from a total of 98 different source countries between 2010 and 2015 (inclusive of zero reporting). The monthly level case counts were aggregated by country of infection (as source country) and the reporting country in Europe (as destination country). We transform the absolute count data into a binary response variable, that indicates whether there was an imported case of dengue (1) or not (0) in a destination country, in a month.

### Air passenger’s data

Comprehensive air passengers travel data for 2010 – 2015, was obtained from the International Air Transport Association (IATA)^26^. The data included true origin, connecting points and final-destination airports for all routes in the world and their corresponding passengers’ volume. The data contains over 11,996 airports in 229 different countries and their territorial dependencies. The passengers’ travel volume for each route worldwide was available at the country level and at a monthly timescale. This data was used to construct a monthly passenger flow from all countries worldwide with a final destination in Europe (accounting for all connecting flights) between 2010 and 2015. The data also included the passengers’ flow between European countries.

### Connectivity indices between a source and destination country

Drawing on the underlining concept of spatial interaction modelling, that inflow between two locations is a function of the attributes of the source and destination and their corresponding interaction^27^. Connectivity indices between a source country and a destination country in Europe were previously developed^13^, using different factors that potentially mediate dengue importation risk. The indices were decomposed into components describing the source strength’ (the risk of dengue infection) and the transport or importation potential (the connection between a source country and a potential destination country in Europe). Source strength for all indices was modelled to represent the endemicity of dengue in a source country. While transport and importation potential were modelled to characterize seasonal dengue activity, incidence rates, geographical proximity, epidemic vulnerability, air passenger volume, population size and wealth of a source country as mediating risk factors. The connectivity indices and their descriptions are listed in Table 1.

**Table 1.** Descriptions of the variables in the dataset.

| Dataset | Variable name | Description | Data source |
| --- | --- | --- | --- |
| Connectivity indices | Dengue activity <sup>(1)</sup> | Notification of one or more confirmed cases in the given month (January-December) in the source country. | HealthMap<sup>30</sup> |
|  | Dengue seasonality <sup>(2)</sup> | Notification of one or more confirmed cases (dengue activity) in a given month, for two or more years from 2010 through 2015, in the source country. | HealthMap<sup>30</sup> |
|  | Dengue Incidence | Annual dengue incidence estimates of the source country. | IHME<sup>31</sup> |
|  | Geo Distance | Geographical (great circle) distance between centroids of the source country and destination country (spatial connectivity). | CEPII<sup>32</sup> |
|  | Vulnerability index | The epidemic vulnerability of the source country. | RAND<sup>33</sup> |
|  | Source GDP | Gross domestic product (wealth) of the source country. | World Bank <sup>34</sup> |
|  | Source Population | The total population of the source country. | World Bank <sup>34</sup> |
|  | Arriving Pax | Total air passengers from source country to a destination country in Europe. | IATA<sup>26</sup> |
| Centrality measures <sup>(3)</sup> | Degree | The number of links or connections that a node has. | Calc. <sup>(4)</sup> |
|  | Betweenness | The number of times a node lies on the shortest path (geodesics) between other nodes in the network. | Calc. |
|  | Closeness | The average number of steps required to access every other node from a given node. | Calc. |
|  | Eigenvector | The combined measure of how many connections a node has (i.e. its degree) and the centrality of the other nodes that it is connected to. | Calc. |
| Created variable | Time effect <sup>(5)</sup> | Set of dummy variables to account for time effects (monthly timescale). |  |

<sup>(1)</sup> Coded as a binary variable, indicating an activity (1) or not (0); <sup>(2)</sup> Coded as a binary variable, indicating a seasonal pattern (1) or not (0); <sup>(3)</sup> Centrality measures for source and destination countries were added as separate variables in the dataset; <sup>(4)</sup> Calc= calculated from the air passengers’ data; <sup>(5)</sup> Set of 72 dummy variables, coded as 1 for a given month and 0 for all other months. Detailed mathematical equations for all variables can be found in Salami et al<sup>13</sup>.

### Centrality measures of the air transport network

Using the monthly air passenger’s data, we constructed a weighted directed network. The network for each month was denoted by *G*_*m*_ = (*V, E*), where *V*_*G*_ is a set containing all the nodes (or vertices), while *E*_*G*_ contains all the edges, with *m* indicating the month (m = 1, 2, 3… 72, covering the years of 2010 – 2015). Nodes represented all countries worldwide, while edges represent the flow of passengers from a source country to a destination country in Europe. Four different centrality measures were used to analyse the network and quantify the capacity of a source node to influence transportation of dengue or a destination node’s propensity to receive an imported case of dengue, by virtue of their connection topology within the network. Centrality measures and their descriptions are listed in Table 1.

### Feature (variable) engineering

One fundamental step in building machine learning models is the process of feature engineering, i.e. using domain-specific knowledge to create new features (i.e. variables) or transform and encode existing original data into a more informative format^28,29^. For this analysis, we created an additional variable based on our a priori knowledge of the data. The reporting rate for dengue data was heterogeneous across the EU/EEA member countries, as some countries were not consistent in monitoring and reporting to the ECDC (we considered “zero reporting”, which designate that no imported case was recorded during the reporting period, but a report was submitted to the ECDC). Hence to account for this variability in reporting rates, we created a time effect variable – coded as a set of dummy variables, 1 for a given month and 0 for all other months. This variable not only controls for reporting rates, but for other potential time-specific effects that might be restricted to a given period. Time events that might increase or decrease passengers’ traffic to a specific country, and in turn affect dengue importation (e.g. the introduction and/or discontinuation of an airline carrier or route).

### Data pre-processing and splitting

The dataset used to build our machine learning models consists of the connectivity indices and centrality measures of the air transport network. The single unit of analysis is a source-destination country pair, at a monthly timescale, and a binary response variable coded to indicate an imported dengue case (1) or not (0).

Before model training, we performed the following pre-processing analyses to the full dataset. First, we examine the correlation between our predictor variables, by using Spearman’s correlation to rank the statistical dependencies. Several pairs of continuous variables displayed moderate-to-high pairwise correlations. Figure 1 shows the correlation matrix between the continuous predictors in our dataset. Most of the centrality measures (for source and destination countries) were highly correlated, example the betweenness and eigenvector centrality for source countries had a Spearman’s *ρ* = 0.99 This is not unusual, as on average, centrality measures are highly correlated in a network^35^. The highly correlated pairs are practically redundant in conventional regression modelling and the heuristic approach to dealing with this is to exclude one. However, in our case, we did not manually exclude variables, as we focus on the predictive power of the entire bundle of variables as oppose the estimated coefficients of individual variables. Likewise, the suite of algorithms compared, each utilizes a combination of inbuilt feature selection and penalization functions to exclude redundant variables in their ensembling and mitigate the effect of multicollinearity.

**Figure 1.**
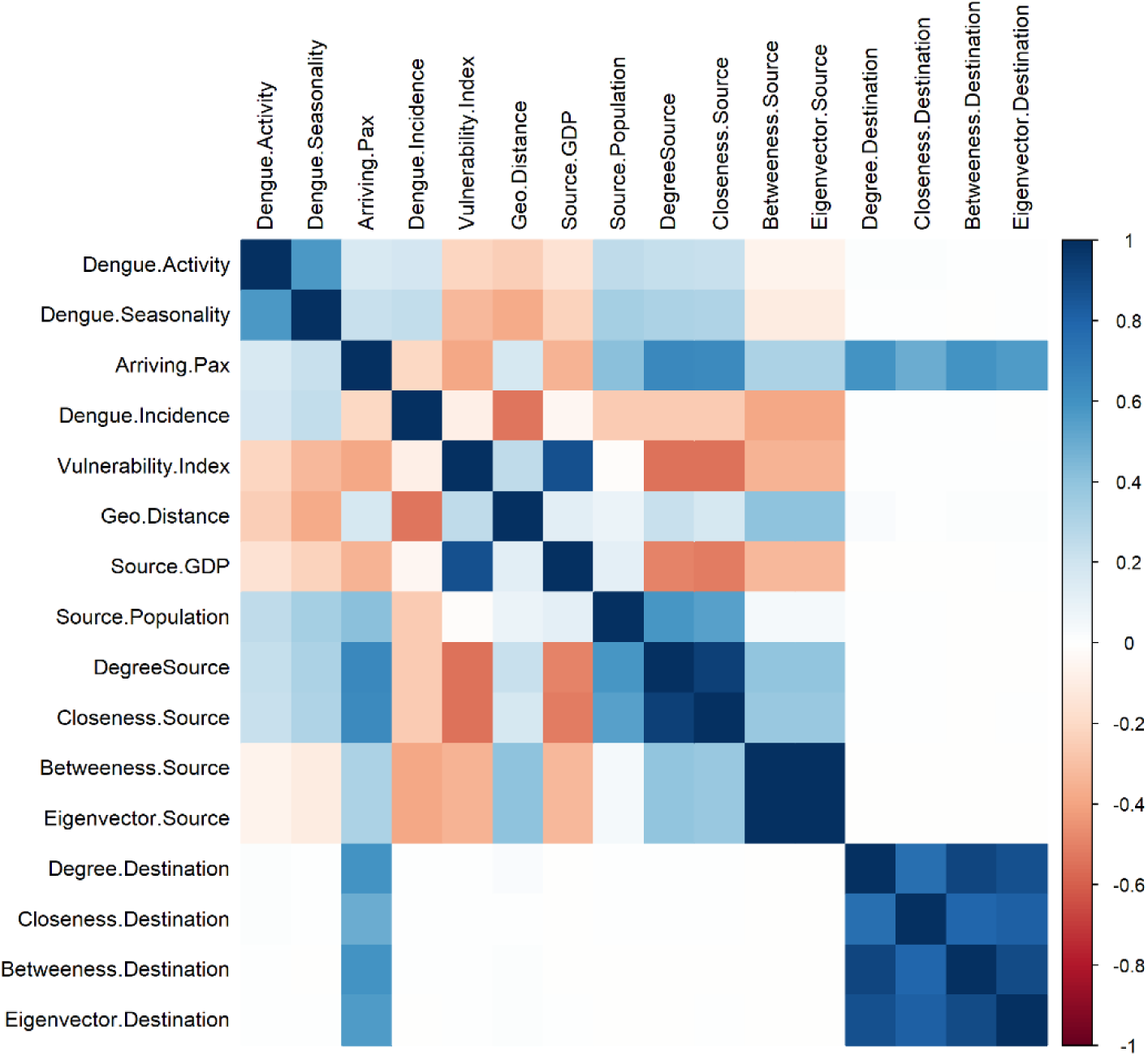
Spearman correlation matrix of continuous variables. Correlation is computed from the full dataset and coloured according to magnitude. Blue colours indicate strong positive correlations, red indicates strong negative correlations, and white implies no empirical relationship between the variables. Figure was generated using R programming language version 3.6.1^39^.

Next, we randomly split the dataset into two sets, 70% into a training subset and 30% into a testing subset. This split was done based on the distribution of our outcome variable (i.e. binary response of an imported case of dengue (1) or not (0)), with sampling occurring within each category, thereby preserving the overall class distribution of our data. Our full dataset contained a total of 2, 055 unique country pairs (i.e. source -destination country pair), with a corresponding total of 147,960 monthly observations. An imported case was recorded in 1,937 observations, i.e. 1% of the total observations had an imported case. These observations were split into 1366 for the training subset and 573 for the testing subset, at a 70:30 ratio (a similar split was done for the observations with no imported cases). The training subset is used to build and tune the various machine learning models, while the test subset is used to evaluate the predictive performance of the models.

Due to the imbalance class distribution of the outcome variable we model this as a rare event, using a post hoc sampling approach to attenuate the effects of the imbalance during model training^36^. The synthetic minority over-sampling technique (SMOTE), was used to subsample the training subset data to create a roughly equal distribution within the classes. SMOTE utilizes a hybrid of either up-sampling, to synthesize new data points in the minority class or down-sampling, to down-size the majority class^37^. Our training dataset was balanced to a 3:4 ratio of an imported case to no imported case. The testing subset was maintained to reflect the original imbalance as a quality assurance of the predictive model performance. Lastly, before training, we apply a data transformation on all continuous variables in the dataset, by centring and scaling them. This transformation ensures that the variables have a zero mean and a common standard deviation of one, thereby improving normality and the numerical stability of the model calculations^38^.

### Model selection

Our training model is a classification-based model, to predict the probability that a case of dengue is imported into a country in Europe. There are several classification techniques (or classifiers) employed in machine learning models. The choice of the suite of algorithms we tested, was a trade-off between, meta-algorithm that fits our classification problem and those with built-in feature selection. Other algorithmic and systematic features that were considered include regularization (to handle the effects of multicollinearity), hyperparameter optimization (model tuning capabilities), and efficient computation time. To build our predictive model, we compare four widely used classifiers algorithms in machine learning, as listed below:

Partial least squares (*pls*) implements a supervised version of principal component analysis, using a dimension reduction technique. This technique first summarizes the original variables into a few new variables called principal components (PCs), as supervised by their relationship to the outcome variable. These components are then used to fit a linear regression model^40^. For classification problems, the partial least squares discriminant analysis variant is fitted. This method has an embedded feature selection and regularization^41^.

Lasso and elastic-net regularized generalized linear models (*glmnet*) implement a logistic generalized linear model via penalized maximum likelihood. The addition of a penalty shrinks the coefficients of the less contributive variables toward zero (L2 ridge penalty) or absolute zero (L1-Lasso penalty)^42^. The *glmnet* implements a combination of both L1 & L2 penalties (otherwise called elastic net penalty), for its regularization and simultaneous feature selection.

Random forest (*randomForest*) is a bootstrap aggregated (or bagged) decision tree-based ensemble technique. The algorithm constructs multiple decision trees by repeat resampling of the training dataset and outputs the mode of the classes as a consensus prediction. The trees are created independently from a random vector distribution; hence each individual tree is heterogeneous with high variance and casts a unit vote for the most popular class^43^. By averaging several decision trees, it intuitively avoids overfitting and performs an embedded feature selection.

Extreme gradient boosting (*xgboost*) implementation of a gradient boosted decision trees ensemble technique. The gradient boosting framework iteratively refines its model, to create a strong classifier by combining multiple weak classifiers in a stage-wise manner to minimize the loss function^44,45^. The *xgboost* algorithm is a commonly preferred classifier, because it utilizes parallelization and distributed computation for implementation, thereby ensuring high efficiency in computation time and resources^45-47^.

### Model tuning and validation

Machine learning models can be prone to overfitting, to mitigate this we implemented a model building approach that encompasses model tuning and repeated evaluation during training. We use a methodological resampling technique of the training dataset, i.e. five repeats of 10-fold cross-validation (CV). The 10-fold CV randomly partitions the training dataset into 10 sets of roughly equal size, one set retained, and the others used to fit a model. The retained set is used to estimate model performance. The first set is then returned to the training set and the procedure iterated until each set has been used for validation. This whole process is repeated five times before results are aggregated and summarized. This procedure automatically chooses tuning parameters associated with optimal model performance.

Candidate models were evaluated using the following performance metrics: area under the receiving operating characteristic curve (AUC), sensitivity (true positive rate), and specificity (false positive rate). Our final candidate model was selected based on the receiving operating characteristic curve (ROC) threshold, which maximizes the trade-off between sensitivity and specificity^48^. The ROC curve evaluates the class probabilities across a continuum of thresholds, with an arbitrary (algorithmically set) “optimal” cut point for determining what percentage of probability is accepted in classifying an imported case of dengue.

### Model interpretability

We utilized the model-agnostic approach to provide interpretation of our optimal model. Model-agnostic methods work by extracting post-hoc explanations from an original machine learning model^23^. This involves training an interpretable model on the predictions of the original model^49^ and/or by changing the inputs of the original model and measure the changes in the prediction output^50^. We employ the use of recent model-agnostic tools^51,52^with both global and local scale interpretability functions. Global interpretation helps to understand the modelled relationship and distribution of the predicted target outcome (i.e. dengue importation) based on the input variables, while local interpretation zooms in, to help understand model predictions for a single instance (i.e. a single unit of observation or analysis).

We obtained global interpretations of our final candidate model through the following, variable importance, and partial dependence plots^49,53^. Variable importance measures the contribution of each input variable, by calculating the increase in the model’s prediction error after permuting the variable^54^. While the Partial dependence plots (PDP) are graphical renderings of the prediction function that helps visualize the relationship between the variables and predicted outcome^46,55,56^. The relative importance of each variable is normalized to have a maximum value of 100, with higher scores indicating the most influential variable. We note that it may not be feasible to explore in detail the relationship of all variables in our model. Hence, we set an arbitrary cut-off on the variable importance measures at a value >50, to determine a subset of variables to focus on.

Local interpretation of our model was implemented via the use of local surrogate models, otherwise called-Local interpretable model-agnostic explanations (LIME)^23,50^. The underlining assumption of LIME is that complex black box models are linear on a local scale, hence a simple (surrogate) model can be fitted for an individual observation that mimics the behaviour of the global model at this locality. The simple model and its variable weights are then used to explain the individual predictions locally. To demonstrate the LIME technique, we selected 10 single observations from our initial testing subset. These observations were sampled methodologically to include both classes (i.e. imported case [1] or not [0]) and representative of countries with a high and low frequency of dengue importation. We set the number of variables to best describe the predicted outcome, as the 5 most influential. The resulting weights for these variables are plotted to explain the local behaviour of the model. The plots delineate if a variable increase or decreases the predicted probability of an imported case of dengue (detailed vignette for the LIME techniques can be found here^57,58^.

### Statistical software

All statistical analyses were performed with R Programming Language version 3.6.1^39^. For uniformity in our model build, we utilized the classification and regression training (*caret*) R package, this is an interface to a vast amount of available machine learning algorithms^38^. The package streamlines the process of building and validating predictive models by using a set of intuitive call functions. Supporting packages for specific functions includes: *pls*^*41*^, *glmnet*^*42*^, *randomForest*^*59*^, *xgboost*^*60*^, *plyr*^*61*^, *doSNOW*^*62*^, *DMwR*^*63*^, *pROC*^*64*^, *pdp*^*56*^, *iml*^*51*^, *lime*^*52*^ and their various dependencies.

## Results

### Model prediction performance

We compared the prediction performance of the different classifiers’ algorithms, in their ability to predict an imported case of dengue. Models were evaluated with the testing dataset, utilizing the area under the receiver operating characteristic curve (AUC) as the quantitative measure for performance comparisons. All four models performed comparably well, with AUC scores above 0.80 (Table 2). AUC score using *pls* was 0.88 (95% CI, 0.86 to 0.90); *glmnet* was 0.89 (95% CI, 0.87 to 0.91); *randomForest* was 0.97 (95% CI, 0.96 to 0.98); and *xgboost* was 0.97 (95% CI, 0.96 to 0.98). Performance metrics for each model are depicted in Table 2. Figure 2 also shows the ROC curve plots for the different models.

**Table 2.**
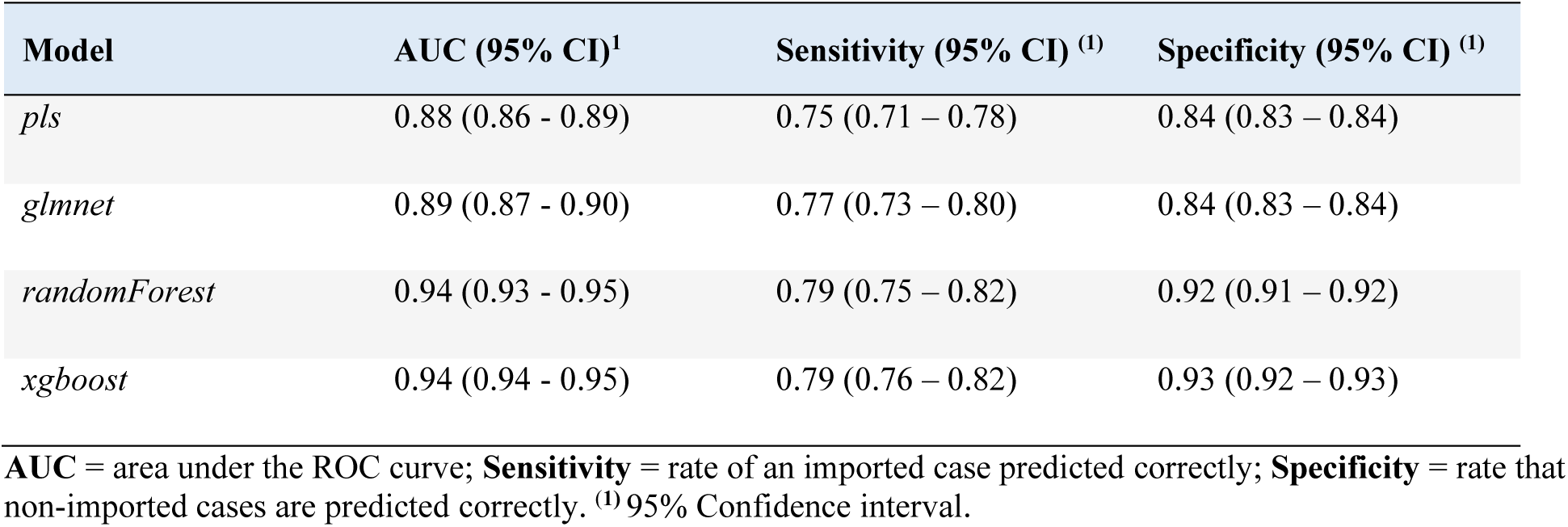
Comparison of the prediction performance of the different models.

**Figure 2.**
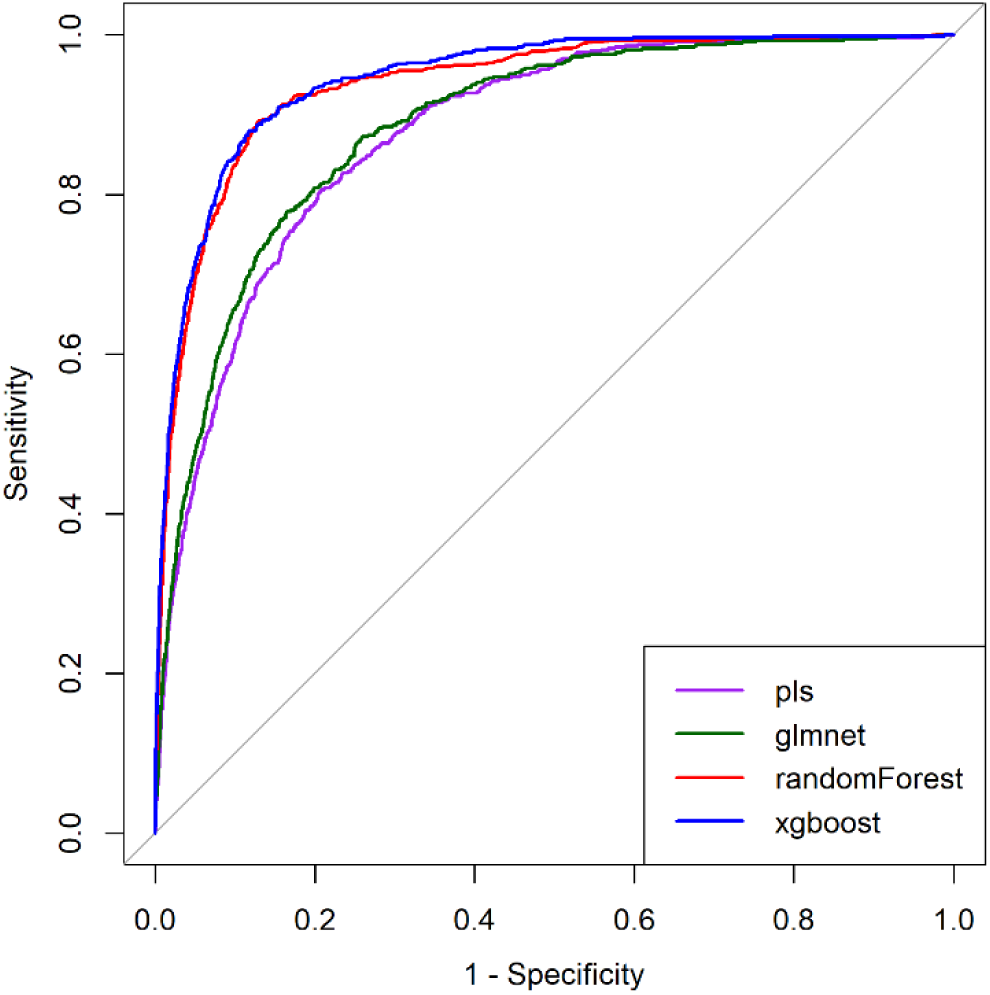
Comparison of the receiver operator characteristic (ROC) curve for the different models. Curves characterize the trade-off between the sensitivity (true positive rate) and 1minus specificity (false positive rate). The *y*-axis = sensitivity and the *x*-axis = 1 minus specificity.

The AUC score indicates that predictions from the *randomForest* and *xgboost* models were better fitted to the dataset, outperforming the *pls* and *glmnet* models (with the pls being the least fitted). The *randomForest* and *xgboost* models had similar performance across the metrics with nearly negligible differences (Table 2). However, they had a distinction in their ROC curves effective threshold (Figure 3). The ROC threshold appropriately maximizes the trade-off between sensitivity and specificity. With the best threshold cut-off at 68% (i.e., only probabilities greater than 0.68 were classified as an imported case of dengue, Figure 3a) the *xgboost* model outperforms the *randomForest*, (cut-off at 0.64, Figure 3b) in a competitive comparison of prediction accuracy. With a true positive rate of 0.88 and a false positive rate of 0.12, the *xgboost* model was selected as the optimal model for our dataset. Hence, our final predictive model was able to predict 88% of dengue importation cases in our test dataset accurately (Figure 3a).

**Figure 3.**
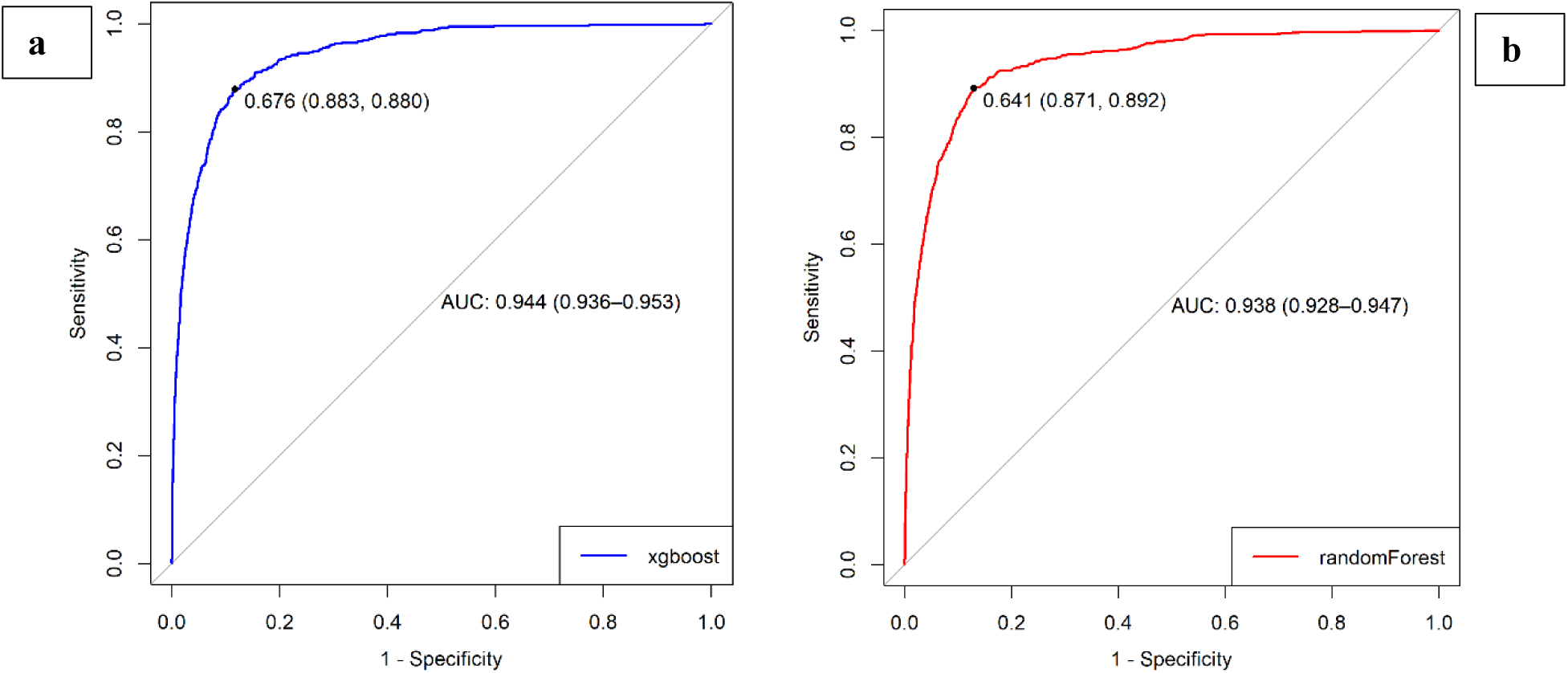
Comparison of the receiver operator characteristic (ROC) curves for extreme gradient boosting and random forest models. The dot on both plots indicates the value corresponding to the “best” cut-off point threshold for each model that appropriately maximizes the trade-off between sensitivity and specificity. The numbers in parentheses are (specificity, sensitivity). Extreme gradient boosting (a) cut-off was at 68% (i.e., probabilities greater than 0.68 are classified as an imported case of dengue), delivering a specificity of 0.883, sensitivity of 0.880, while random forest (b) cut-off was at 64%.

### Model interpretability

The best performing model (i.e. *xgboost*), was examined further for interpretation. Figure 4 illustrates the variable importance scores for the10 most influential variables, from our optimal (i.e. *xgboost)* model. With an arbitrary cut-off at >0.50, the subset of our ‘most important’ variables included the following: Source country’s dengue incidence rate; population; the number of arriving passengers; betweenness, closeness and degree centrality measures of the destination country. Figure 5 illustrates a visual representation of the relationship between this subset of variables and the predicted response while accounting for the average effect of the other predictors in the model. These plots demonstrate that the probability of an imported case of dengue increases on average for source countries with higher incidence rates, large population size and higher passenger traffic. Likewise, the probability increases for destination countries with higher betweenness, closeness and degree centrality measures.

**Figure 4.**
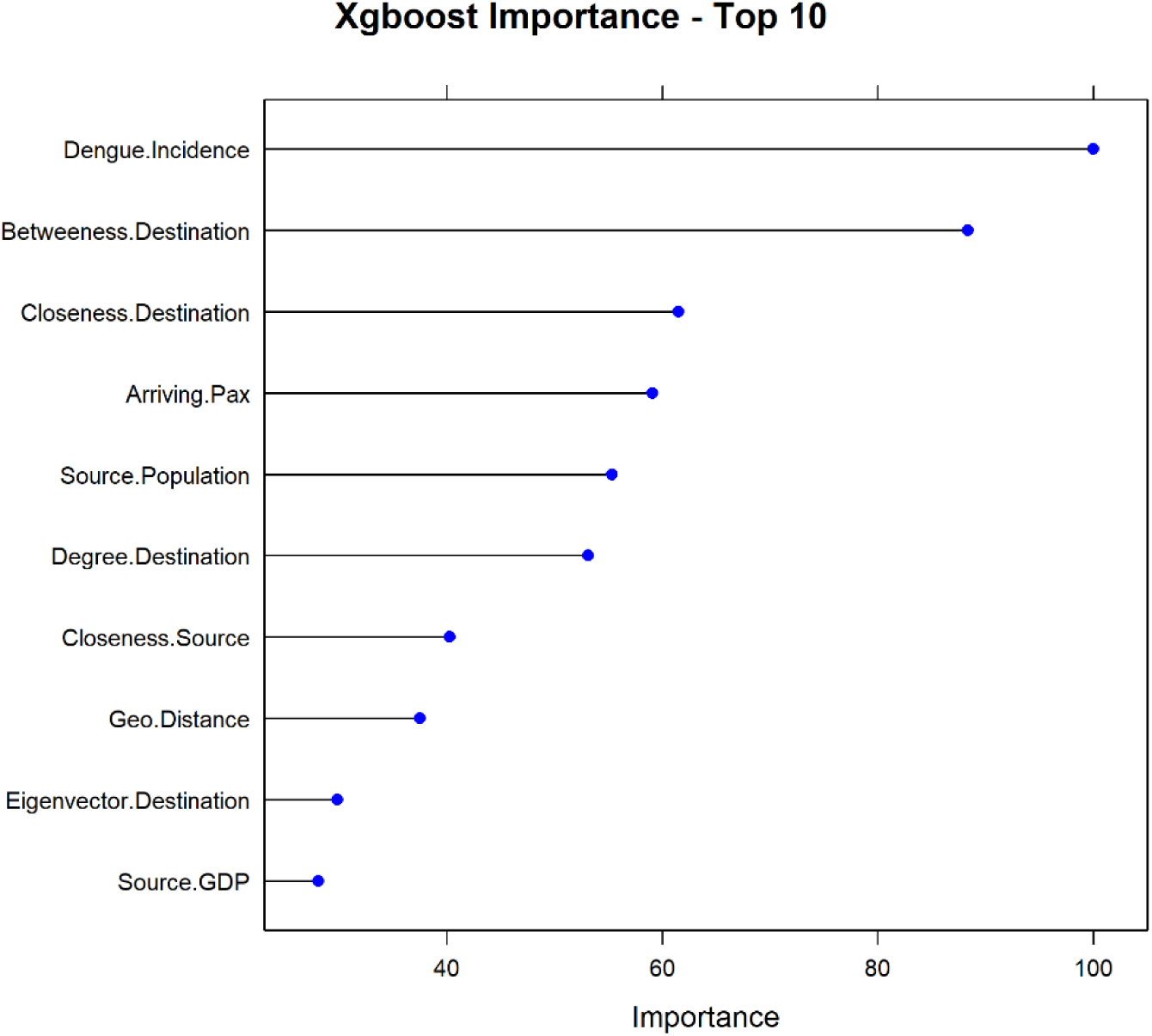
Variable importance plots. Top 10 most influential variables from the extreme gradient boosting model. The relative importance of each variable is normalized to have a maximum value of 100, with higher scores indicating the most influential variable.

**Figure 5.**
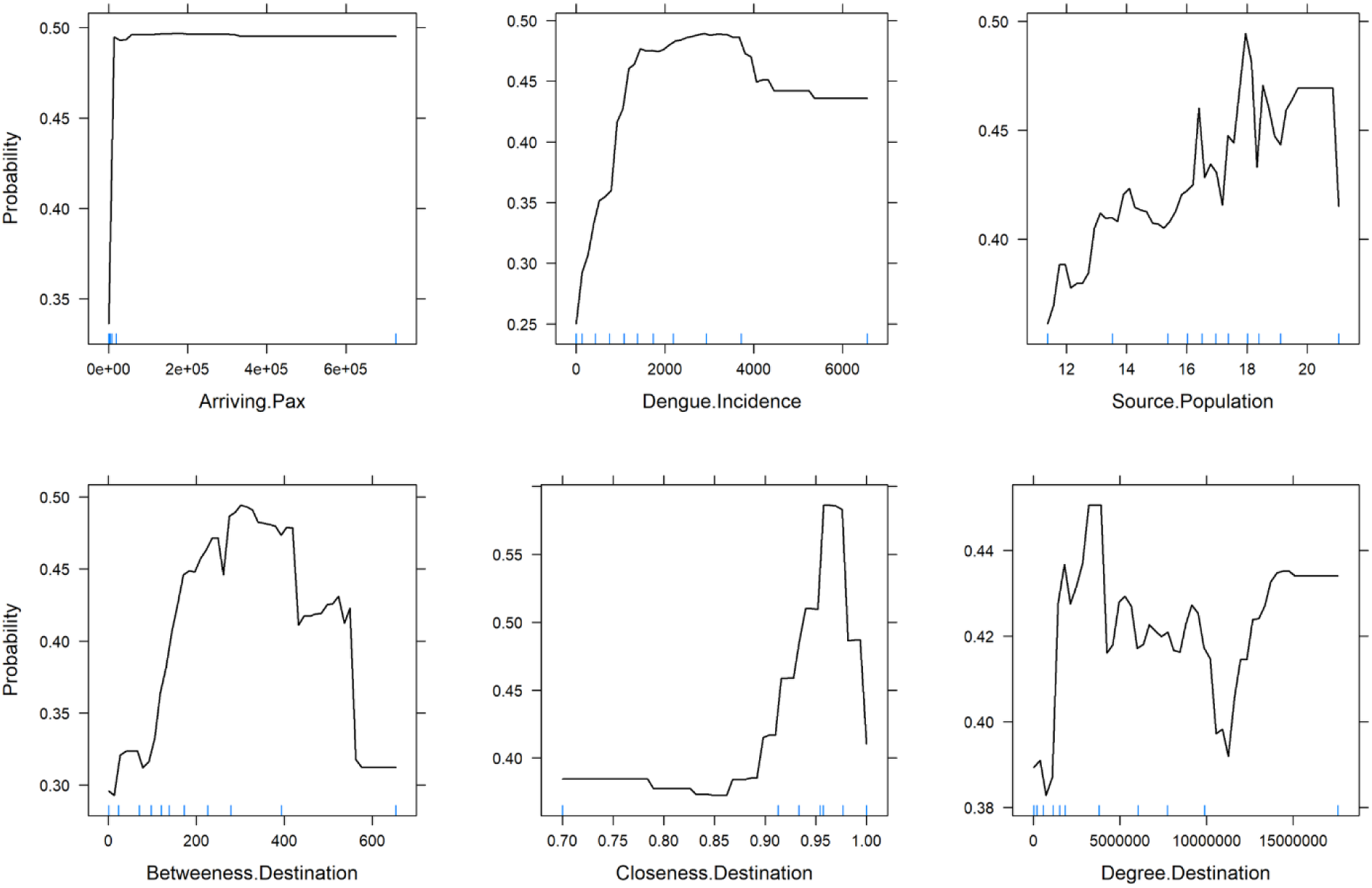
Partial dependence plots for a sub-set of the most influential variables in the optimal model predicting the probability of an imported case of dengue. The optimal model is the *xgboost* model. Sub-set variables represent variables with a variable importance ranking score >50. *Y*-axis is set on a probability scale since our model was a classification model; Blue rug marks at the inside bottom of plots indicate the min/max and deciles of the variable distribution. Top plots show the 3 most influential connectivity indices, while bottom plots show the 3 most influential network centrality measures.

Note: to guard against over-interpreting the partial responses, we added rug displays to the plots, i.e. tick marks indicating the minimum, maximum and deciles of the variable distribution. Our interpretation of the partial response was limited to the regions within the minimum and maximum.

In addition to providing global explanations of our optimal model, we provide a local explanation for individual predictions given for 10 single observations, i.e. a single unit of analyses, source– destination-month combination (Table 3). Figure 6 gives a visual representation of the first four single observations in local subset data, each plot shows the predicted probability of each observation, being an imported case of dengue. Likewise, it shows the five most influential variables that best explain the model’s prediction at the local region of the single observation, and whether these variables increase (supports) or decrease (contradicts) the probability of an imported case of dengue. With these results, we can infer that for case 1 (i.e. Indonesia-to-Germany, for February 2010), the local model-predicted probability of being an imported case, was 94%. The top five variables influencing this probability were: closeness and betweenness centrality measures of Germany, the incidence rate of dengue and closeness centrality measures of Indonesia and the geographical distance between both countries. Conversely, case 3 (i.e. Tanzania to the United Kingdom, for December 2014), had a similar set of variables as most influential, however, the incidence rate of Tanzania decreases the probability of having an imported case. This demonstrates how variables influencing predictions for a single observation can differ at the local scale. Finally, to ensure the trustworthiness of the local model, we compared the predicted probability of the local model to that of the global optimal model for each observation. There was no difference in the predicted probabilities of the global and local model (analytical comparison not shown). Overall, the local interpretation provides insights into the variations of the individual predictions and provides an important aspect to assuring trust of the model.

**Table 3.** Ten selected individual observations for LIME model. (unit of source–destination-month combination).

| Case # | Source country | Destination Country | Month<sup>(1)</sup> | True Class<sup>(2)</sup> | Predicted Class<sup>(3)</sup> | Probability<sup>(4)</sup> | Top Five variables from LIME<sup>(5)</sup> |
| --- | --- | --- | --- | --- | --- | --- | --- |
| 1 | Indonesia | Germany | 2 | 1 | 1 | 0.94 | Supports: Closeness. Destination; Geo Distance; Betweenness. Destination; Closeness. Source; Dengue Incidence |
| 2 | Brazil | Norway | 61 | 1 | 1 | 0.80 | Supports: Closeness. Destination; Geo Distance; Source GDP; Dengue Incidence<br>Contradicts: Betweenness. Destination |
| 3 | Tanzania | United Kingdom | 60 | 1 | 1 | 0.91 | Supports: Closeness. Destination; Geo Distance; Betweenness. Destination; Source Population<br>Contradicts: Dengue Incidence |
| 4 | India | Sweden | 39 | 1 | 1 | 0.92 | Supports: Closeness. Destination; Geo Distance; Source GDP; Dengue Incidence<br>Contradicts: Betweenness. Destination |
| 5 | Thailand | Italy | 34 | 1 | 1 | 0.90 | Supports: Closeness. Destination; Geo Distance; Betweenness. Destination; Source GDP<br>Contradicts: Dengue Incidence |
| 6 | Vietnam | France | 20 | 1 | 1 | 0.94 | Supports: Closeness. Destination; Geo Distance; Source Population; Source GDP<br>Contradicts: Dengue Incidence |
| 7 | Brazil | Portugal | 43 | 0 | 0 | 0.53 | Supports: Geo Distance; Dengue Incidence<br>Contradicts: Closeness. Destination; Betweenness. Destination; Source GDP |
| 8 | Columbia | Spain | 14 | 0 | 0 | 0.52 | Supports: Closeness. Destination; Geo Distance; Betweenness. Destination; Source GDP<br>Contradicts: Dengue Incidence |
| 9 | Philippines | Austria | 47 | 0 | 1 | 0.77 | Supports: Closeness. Destination; Geo Distance; Betweenness. Destination; Source GDP; Source Population |
| 10 | Venezuela | Ireland | 66 | 0 | 0 | 0.13 | Supports: Betweenness. Destination; Dengue Incidence; Closeness. Source<br>Contradicts: Closeness. Destination; Geo Distance |

<sup>(1)</sup> Month case was reported in the destination country, 1 – 72 months covering the years of 2010 – 2015, e.g. month 1 = January 2010; <sup>(2)</sup> Original classifications of the case, in the test dataset, imported case [1] or not [0]; <sup>(3)</sup> Predicted class in LIME model, imported case [1] or not [0]; <sup>(4)</sup> Prediction probability of an imported case of dengue from LIME model; <sup>(5)</sup> Top five most influential variables, delineated by if variable increases (supports) or decreases (contradicts) the probability of an imported case of dengue.

**Figure 6.**
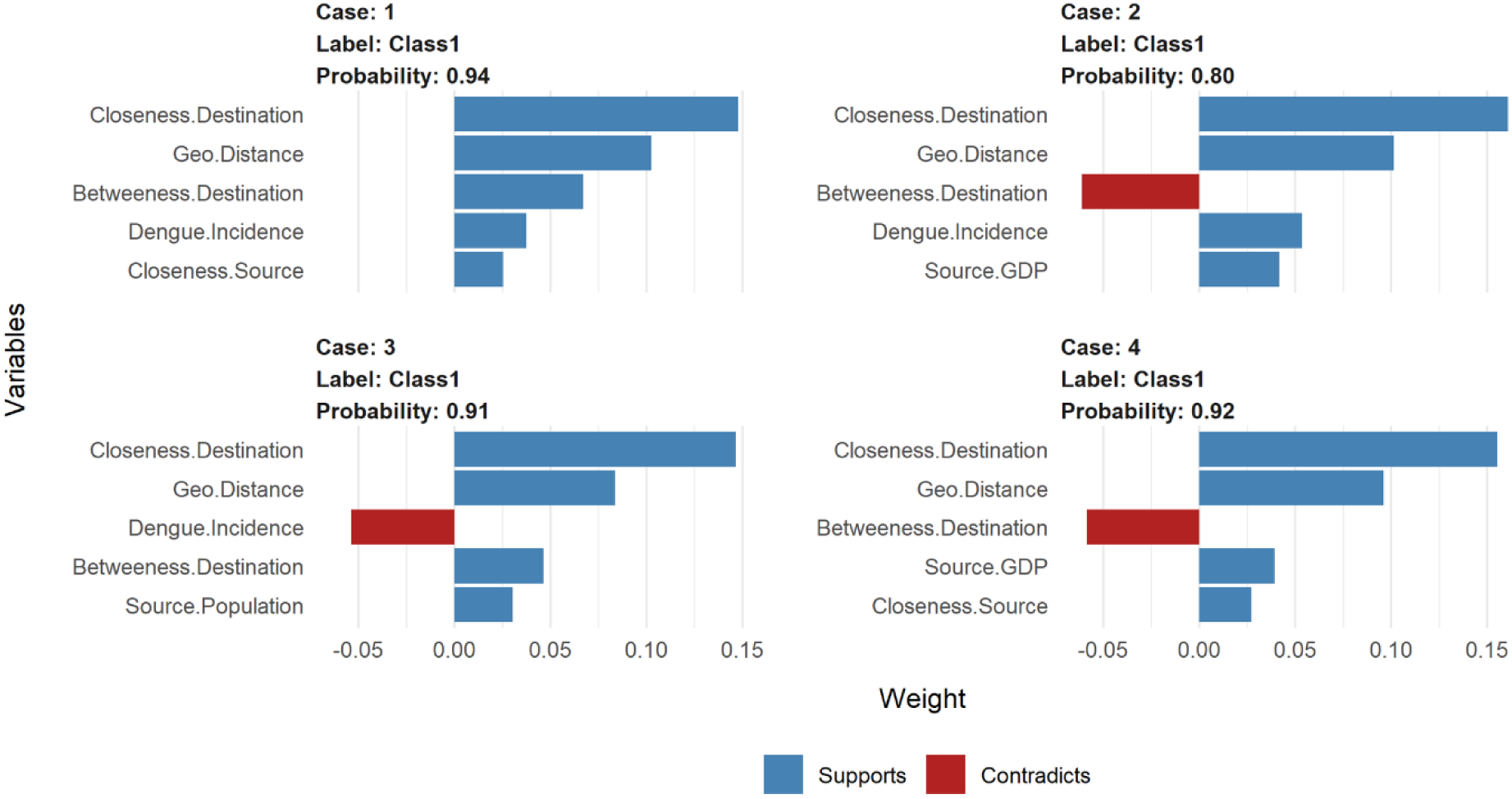
LIME model plots explaining individual predictions. Cases 1-4, as presented in Table 3. Each plot shows the 5 most influential variables that best explain the prediction in the local region. The blue bars represent variables that increase the predicted probability (supports), the red bars represent variables that decrease the probability (contradicts).

## Discussion

Our study demonstrates the use of machine learning modelling approach to predict the probability of having an imported case of dengue in Europe. Using historical dengue importation data, we trained and evaluated four machine learning classifiers algorithms, to develop an optimal predictive model. Our best-performing model was the extreme gradient boosting model with an AUC score of 0.94. Our choice of best performing model was not just based on the AUC results, as this score does not necessarily guarantee the best classifier. Given that our prediction target is the probability of having an imported case of dengue, we expected that our final model performs better in classifying the true positive cases (i.e. maximizing sensitivity). This was achieved by the effective probability threshold of the ROC curve, which offered a trade-off between sensitivity and specificity. Utilizing this we were able to maximize the sensitivity of our optimal model, to correctly predict the probability of an imported case of dengue with an 88% accuracy rate. As a first attempt to train a machine learning model for dengue importation in Europe, we can safely state that our model provides a benchmarking result for predictive performance. Given this predictive performance, our model holds great potential as a forecasting tool which can markedly improve dengue surveillance in Europe.

A limitation of other previous machine learning models is that they deliver high predictive accuracy without explaining why certain predictions are made^14,65^. In this paper, we posit for both accuracy and interpretability of our final model. We focus on demonstrating the practical explanation of our model predictions using recent model-agnostic approaches^23^. Our model included 17 predictor variables, which broadly captures the importation risk factors (as presented by the connectivity indices) and the influence of the air transport network (i.e. the centrality measures). These variables were chosen to reflect the factors known or hypothesized to be relevant to the importation dynamics. So firstly, we provided an overall quantification of the relationship between our model variables and the predicted outcome, by ranking them in terms of their importance. With a further exploratory analysis (via the PDPs visualizations) concentrating on a sub-set of the most influential variables. On average our model predicted a higher probability of an imported case of dengue, from a source country with high dengue incidence rates, large population, and high air passenger volume. These findings support a priori expectation for these factors to increase the importation risk of dengue and are consistent with other studies^12,13,66^. Also, our model predicts a higher probability of an imported case for destination countries with high connectivity within the network (as measured by degree centrality), with a putative connection hub to other countries (betweenness centrality) and connects to other countries in a relatively short amount of time (closeness centrality). Intuitively, this is expected, given that the network centrality of the destination country was modelled to act as a proxy for a country’s propensity to receive an imported case. Hence, destination countries with higher connectivity within the network have an increased risk of importation. These findings are similar to other previous studies that characterize the role of the air transport network structure in mediating epidemic spread^10,11^ and collaborates the results of our previous work^13^.

The above explanations capture the relative contribution of the input variables in predicting the importation of dengue at an aggregated level for Europe. However, the risk probabilities will differ at the country level due to changes in the dynamic attributes of the different country pairs. For example, the dengue activity or seasonality in a source country can vary relative to time (oblivious to high or low incidence rates). Hence, it will be expected that importation risk based on seasonality will have temporal differences between source countries. Also, the volume of air passengers between country pairs are heterogeneous, which can be largely determined by different monthly traffic flows. Also, the topological profile of the air travel network will differ across countries with changes in passenger volume. These heterogeneities may affect the prediction of dengue importation at the country level, with certain variables supporting or contradicting depending on the country pairs in consideration. Using the local interpretable model-agnostic explanations, we were able to asses which variables are most influential on the predictions at a temporal and country-pair level. As illustrated from the examples in Table 3, the probability risk prediction for different country pairs was increased or decreased by different variable combinations at discrete points in time. For example, the predicted probability of a case of dengue from Indonesia (case 1 in Table 3) and Tanzania (case 3 in Table 3), were similar, but vary in the risk factors mediating this prediction. The lower dengue incidence rates in Tanzania (relative to Indonesia), decreases the probability of an importation, however other variables pose an increased risk for an imported case of dengue. So, the local explanations provide insights into the heterogeneities of importation risk at the different country pairing levels. This type of specific country-pair results can be useful in profiling importation risk from a specific source country or region; similar to the route-level risk assessment discussed by Gardner et al ^10^. This kind of information will be useful in guiding the implementation of targeted surveillance and public health preparedness in destination countries. Also, it can guide policy decisions at the European regional level, on how to effectively appropriate surveillance resources for countries at higher risk of dengue importation.

Overall, our work offers the following contributions. First, to the best of our knowledge, this is a first attempt at applying machine learning algorithms to model the risk of dengue importation into Europe. Second, we implement and demonstrate how to apply model-agnostic approaches for obtaining both aggregate (global) and individual (local) level explanations. Finally, we identify and interpret the factors influencing dengue importation through air travel at a temporal and country-pair level. When combined, these contributions can assist public health practitioners looking to develop a reliable, cost-effective, and scalable early warning surveillance system for dengue importation. Although we mainly focus on the theoretical framework of the model using historical data, the results demonstrate that the model can be applied for real-time prediction, assuming the availability of real-time data. However, there are some limitations to this work, that is worth noting for improvement in future research: (1) Dengue incidence rate for source countries was aggregated at a yearly scale, due to paucity of surveillance data at a similar scale to dengue case data (i.e. monthly). This may have overestimated or underestimated the actual effect of incidence and potentially impact the predicted risk probability from a source country. Our approach compensated for this limitation by the inclusion of the dengue activity and seasonality variables. Even though this does not necessarily capture the variability of a finer scale but serves as a proxy. (2) We only evaluated our prediction as a binary outcome (i.e. the probability of an imported case or not) and not a numeric outcome like other similar models for dengue incidence^14,20^. A numeric outcome prediction can be achieved by modifying our model training approach from a classification model to a regression model. Even though, the additional benefit (if any) of predicting a discrete number versus a probability estimate is subjective. However, we do submit that while our approach could serve as a benchmark, we encourage alternative exploration for improved performance and accuracy.

In conclusion, our study demonstrates the efficient and powerful predictive capabilities of machine learning models in predicting the importation of dengue in Europe. Using historical dengue importation data, connectivity indices and air transport network centrality measures, we trained and evaluated a classification model to predict the probability of an imported case of dengue. Then applying recent model-agnostic interpretability approaches we provided an in-depth explanation of the model’s predictions. With the predictive model and model-agnostic interpretability tools at hand, this can be applied at a regional or country level to develop a forecasting tool for dengue importation. Assuming the availability of real-time data, the methods described in this paper can be explored as a technique for developing a real-time early warning surveillance system for dengue importation.

## Data Availability

The air travel data used in this study, cannot be shared publicly because of a nondisclosure agreement with the International Air Travel Association (IATA). The same data can be purchased for use by any other researcher by contacting the International Air Travel Association (IATA)- Passenger Intelligence Services (PaxIS) (https://www.iata.org/services/statistics/intelligence/paxis/Pages/index.aspx).
The disease (dengue) data are available by request from the European Centre for Disease Prevention and Control (ECDC) (https://www.ecdc.europa.eu/en/publicationsdata/european-surveillance-system-tessy). All other relevant data sources are referenced in the article.

https://www.iata.org/services/statistics/intelligence/paxis/Pages/index.aspx

https://www.ecdc.europa.eu/en/publicationsdata/european-surveillance-system-tessy

## Authors contribution statement

Conceptualization, DS, CS, and CC; Data Curation, DS; Formal Analysis, DS; Methodology, DS and CC; Supervision, CS, MM, and CC; Visualization, DS; Writing – Original Draft Preparation, DS; Writing – Review & Editing, DS, CS, MM, and CC. All authors read and approved the final manuscript.

## Acknowledgments

This work was partially funded by Fundação para a Ciência e a Tecnologia (FCT), Portugal (GHTM – UID/Multi/04413/2013 and Project Warden -PTDC/SAU-PUB/30089/2017). DS has a PhD grant fromFCT, Portugal (PD/BD/128084/2016). CC acknowledges support from FCT (CEECIND/02037/2017, UIDB/00295/2020 and UIDP/00295/2020). We greatly appreciate Dominic Freienstein for his assistant in accessing the international air travel association, passenger intelligence services (IATA-PaxIS) data.

## Data availability

The air travel data used in this study, cannot be shared publicly because of a nondisclosure agreement with the International Air Travel Association (IATA). The same data can be purchased for use by any other researcher by contacting the International Air Travel Association (IATA)-Passenger Intelligence Services (PaxIS) (https://www.iata.org/services/statistics/intelligence/paxis/Pages/index.aspx).

The disease (dengue) data are available by request from the European Centre for Disease Prevention and Control (ECDC) (https://www.ecdc.europa.eu/en/publicationsdata/european-surveillance-system-tessy). All other relevant data sources are referenced in the article.

